# Nurturing care practices for children with developmental disabilities in sub-Saharan Africa: A scoping review protocol

**DOI:** 10.1101/2023.09.08.23295265

**Authors:** Silas Onyango, Margaret Nampijja, Paul Otwate, Nelson Langat, Linda Oloo, Kenneth Okelo, Patricia Kitsao-Wekulo

## Abstract

**Background:** The majority of children with neurodevelopmental disorders (NDDs) reside in low- and middle-income countries (LMICs). NDDs is a public health concern in countries in sub-Saharan Africa (SSA). Nurturing care has been recommended as a pathway for addressing the developmental needs and unlocking the full potential of children, including those with NDDs. However, little information exists on the strategies to support children with NDDs using the Nurturing Care Framework in many SSA Countries. The proposed scoping review aims to identify, explore, and map the literature on nurturing care practices for children with NDDs in SSA. The review will also determine gaps in the provision of nurturing care for children with NDD. Further, the review will highlight the drivers of care as well as the experiences of the caregivers in relation to the care provided.

**Methods:** The review will be implemented in six steps: identification of the research question; identification of relevant studies, selection of studies to be included, charting the data and collating, summarizing, reporting the results, and stakeholder consultation. The selection of the articles will be completed in two parts: a database search followed by a manual search. We will search the following electronic databases: PubMed, ScienceDirect, Scopus, Open Grey and AJOL. All studies published after May 2018 to May 2023 that include relevant terms will be identified and included. The research team will develop a data extraction form for use in capturing relevant information from each of the included studies. A patterning chat that will summarize and analyze the key findings of each article will be created.

**Discussion:** We anticipate the study will provide evidence on the existing nurturing care practices for children with NDDs and unearth gaps in the provision of nurturing care for children with NDD. Key determinants of care and the experiences of the parents/caregivers of children will also be identified. The study will provide key recommendations on interventions to improve the quality of care for children with NDDs. Through this study, awareness of the unmet nurturing care needs of these children will be increased. The evidence generated may assist policymakers and stakeholders in addressing the needs of children with NDDs.

## Background

Globally, it is estimated that over 53 million children currently live with NDDs, and the majority of these children live in LMICs (1). Children with NDDs face deficits in neurological and brain functioning that considerably impair their cognition, communication, mobility, and/or social interaction. NDDs, or childhood disabilities, are a significant cause of poor development in children and continue to be a public health concern in many countries in SSA (2). Nurturing care has been recommended as a pathway for addressing the developmental needs and unlocking the full potential of children, including those with NDDs (3). *Nurturing care* refers to a stable caregiving environment that provides for good health, adequate nutrition, opportunities for early learning, responsive caregiving, and safety and security (4). All children need nurturing care to thrive; however, children with NDDs need more intensive nurturing care strategies not only to survive but also to thrive, as they are at a higher risk of poor developmental outcomes.

With nurturing care, known and modifiable risk factors for developmental disorders, such as poor maternal health-seeking behavior and family violence, can be monitored and effectively mitigated. Children with NDDs are often at risk for maltreatment, violence, and neglect due to unfavorable cultural beliefs about disability, discrimination, and family and community stigma (2). Limited resources in most settings in SSA, where the majority of these children live, further constrain their care as the demands for their needs are always high. Moreover, persistent parental stress and negative feelings associated with having a child with NDDs present a challenge and demand parental care (1). Studies on young children in LMICs have generally excluded those with NDDs, and therefore little is known about the importance of interventions such as nurturing care to support child growth and development.

Nurturing care framework was launched to help many children from LMICs who are at risk of poor development survive and thrive in order to achieve their full potential (4,5). Since its launch, the framework has been implemented across the globe, with SSA countries being the main focus. However, the nurturing care for children with NDDs who require more support has not received strong attention. Also, little information exists on the strategies to support children with NDD using the framework in many SSA countries. The objective of the proposed scoping review is to identify, explore, and map the literature on the nurturing care practices for children with NDDs in SSA. The review will also highlight the existing gaps in the provision of nurturing care for children and determine the drivers of the quality of nurturing care provided. Further, the review will highlight the experiences of caregivers of children with NDD in relation to the care provided. It is anticipated that the evidence generated from this study will inform appropriate interventions or practices that can inform policy on nurturing care for children with NDDs in Kenya and similar settings. This will ensure that children living with disabilities in low-resource settings receive the nurturing care they need to thrive to their full potential.

The research questions are:

1. What are the current nurturing care practices for children with NDD in SSA?
2. What gaps exist in the provision of nurturing care for children with NDD?
3. What are the key determinants/drivers of the quality of nurturing care provided to children with NDD?
4. What are the experiences of parents/caregivers of children with NDD in relation to the nurturing care provided?

**Table 1:**
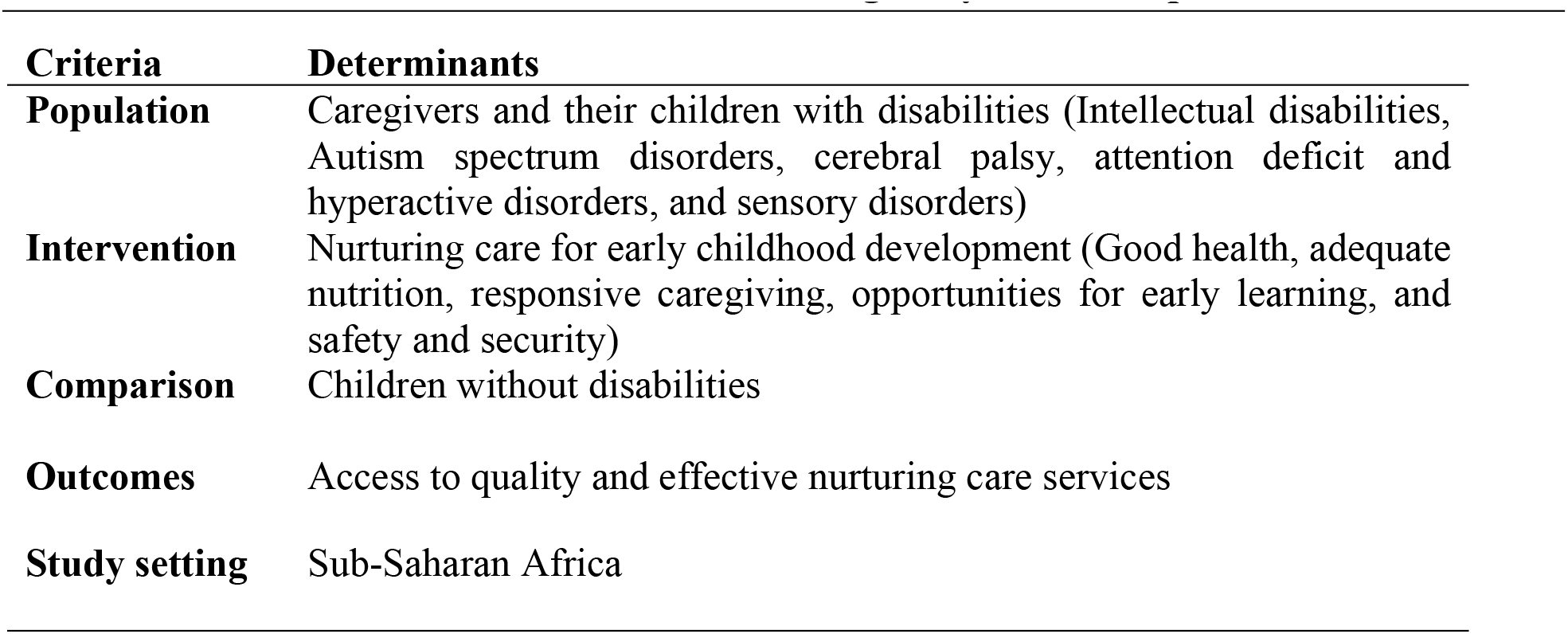
PICO framework for determination of eligibility of review questions Criteria Determinants.

## Methodology

This literature review will utilize the scoping review approach to answer the overarching question, “What are the current nurturing care practices for children with NDDs in SSA?” We chose a scoping review method because it will provide a broader perspective on the existing nurturing care practices across different age ranges. The review will be based on the five components of the nurturing care framework (4) and will follow the proposed methods by Arksey & O’Malley (6) and advanced by Levac and colleagues (7). The scoping review will be implemented in six stages namely; i) identifying the research question, ii) identifying relevant studies, iii) study selection, iv) charting the data, v) collating, summarizing, reporting the results and stakeholder consultation. The selection process will follow the recommendations in the preferred reporting items for systematic reviews and meta-analyses extension for scoping reviews (PRISMA-ScR) checklist (8). The team used excel to manage the review process.

### Identifying relevant studies

The research team will search published and unpublished literature from all the key relevant electronic databases. All studies published after May 2018 to date will be searched using search terms including “developmental disabilities” “nutrition” “health” “nurturing care” “responsive care” “early learning” “safety” “security’ “children with disability” “ADHD” “autism” “cerebral palsy” “learning disorder” “intellectual disorder” “communication disorder” and “sub-Saharan Africa” will be identified and included. In addition, we will search websites such as the World Health Organization (WHO) to identify any potential literature or publications relevant to our study’s main question. Relevant unpublished literature will be identified through a search of conference abstracts, organizational reports, and ProQuest Dissertation & Theses Global. If the search produces too many irrelevant studies, we will consider revising the search strategy to include only relevant terms that could yield reasonable results. The nurturing care framework was launched in May 2018, thus the choice of the period of the scoping review.

**Table 2:**
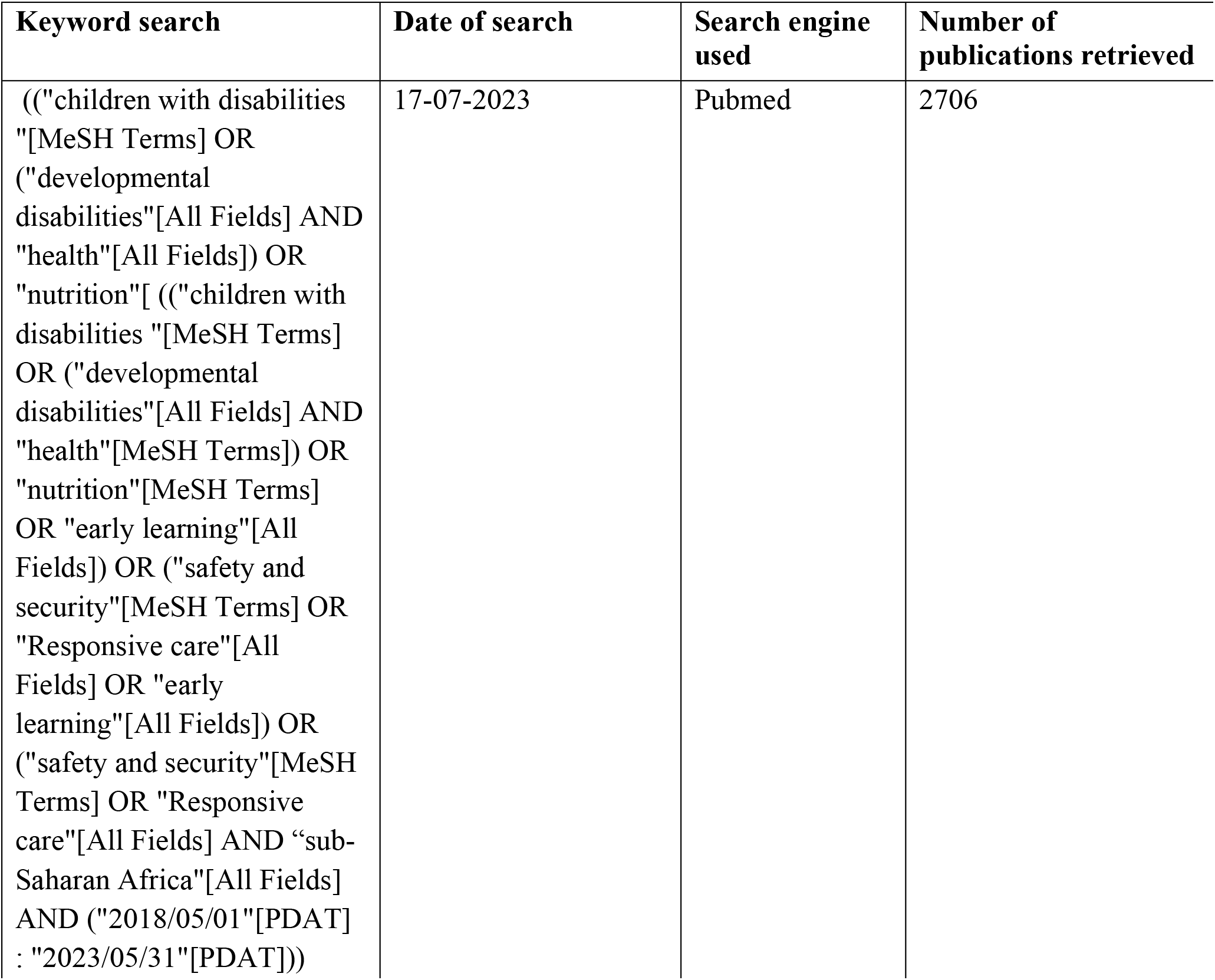
A draft search strategy from PubMed.

### Database searching

The database search for the proposed review will be completed in two parts: one by means of a database search and the other by means of a manual search. Most studies have used database searches to complete the scoping reviews, while manual searches are less common. We proposed a combination of the two search methods. The review team will carry out the reviews independently.

### Systematic manual search

The five components in the nurturing care framework make the topic under review interdisciplinary in nature, and therefore potential articles may not be found in databases or in other journals that may not be in databases. Therefore, it is necessary to use the manual searching method to expand the scope of the search for potential articles. Based on Teare and Taks recommendations (9), a systematic manual search will be conducted in three steps. The first step will be to select top journals based on the five components of nurturing care and their relation to NDDs. The journals will be identified by the impact factor. The research team will then search for articles published after May 2018 that are related to the research topic by keywords. The articles will be selected based on their titles and abstracts, as well as a full-text reading. The articles that meet the criteria are eventually included in the selection. Finally, after selecting the relevant articles in the top journals, the reference lists of these articles will be checked one by one to find other relevant journals, followed by a manual search of these relevant journals. Steps 2 and 3 will be repeated until no new journals appear.

### Systematic database search

The electronic databases to be searched are: PubMed, ScienceDirect, Scopus, Open Grey, and African Journal Online (AJOL). The systematic data search will follow two levels of screening: i) a title and abstract review, and ii) a full-text review. In the first level of review, each of the researchers will independently screen the abstracts and titles against a set of inclusion criteria. In the second level, the researchers will independently assess the full-text articles that made it through the screening stage to ascertain that they meet the inclusion and exclusion criteria. Studies that report participants outside the SSA. Any disagreement in full-text articles will be reviewed a second time by a third reviewer, and further disagreements about eligibility at the full-text review level will be resolved through discussion until consensus is obtained.

**fig. 1.**
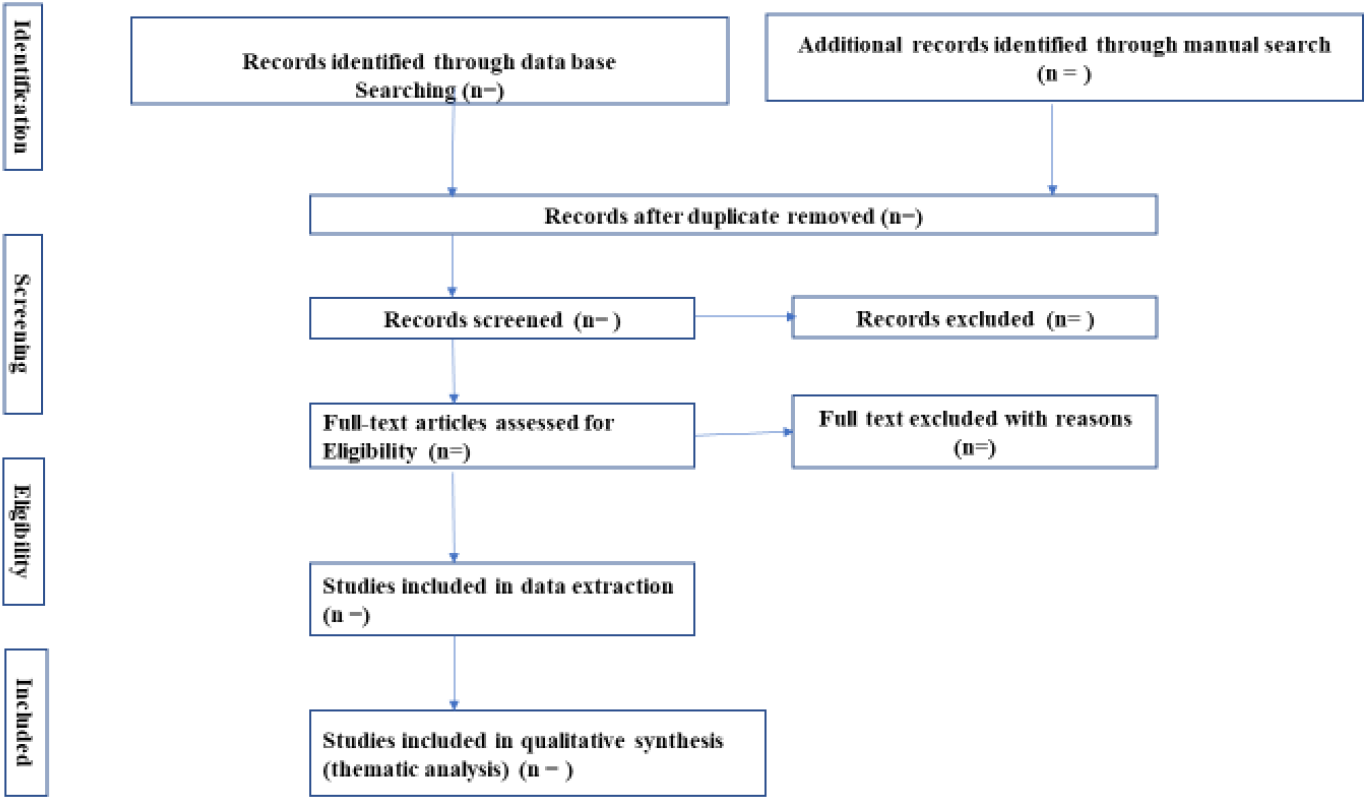
Study selection procedure

### Inclusion and exclusion criteria

The studies that will be included must meet the following criteria: i) they must focus on children ages 0–5 with NDD; ii) they must report any of the five components of the nurturing care framework (health, nutrition, opportunities for early learning, responsive caregiving, and safety and security); iii) they must be published after May 2018, iv) they must include participants from SSA; and v) they must be published in the English language. Any article that will be found relevant by the reviewers and is in line with the research questions will be included in the full review.

### Charting the data

To capture data electronically, the research team will develop a data extraction form that will capture relevant information from each of the included studies. The form will include study identification (using the last name of the first author and year of publication), title, aim, setting, population, sampling method, data collection method, outcomes, data analysis, conclusion, or most relevant findings. The data extraction form will also have a section for comments by the investigators. The form will be reviewed by all investigators before implementation to ensure that it accurately captures important information.

### Collating, summarizing, and reporting the results

We propose to use the updated Patterns, Advances, Gaps, Evidence of Practice, and Research Recommendations (PAGER) framework for the analysis and reporting of data for this review (10). After completion of article searches, the analysis of findings will be undertaken. The first step of analysis will be to create a patterning chat that will summarize and analyze the key findings of each included article, bringing them together into unique key themes based on the five components of the nurturing care framework. We will then describe the progression of these patterns (themes), reflecting the dynamic state of the nurturing care practices for children with NDDs in SSA. The team will then identify any research gaps in the articles, especially in the provision of nurturing care for NDDs. In addition, we will take note of areas that have been extensively explored and do not require further exploration. After identifying the gaps, we will extract from each article information useful to practitioners, academics, and policymakers. Finally, from the identified gaps and practical information provided to key stakeholders, we will suggest research recommendations, including where future research should be focused, gaps in existing research, and areas that have been exhausted. These recommendations will be useful in the design of future research on this topic.

**Table 3:**
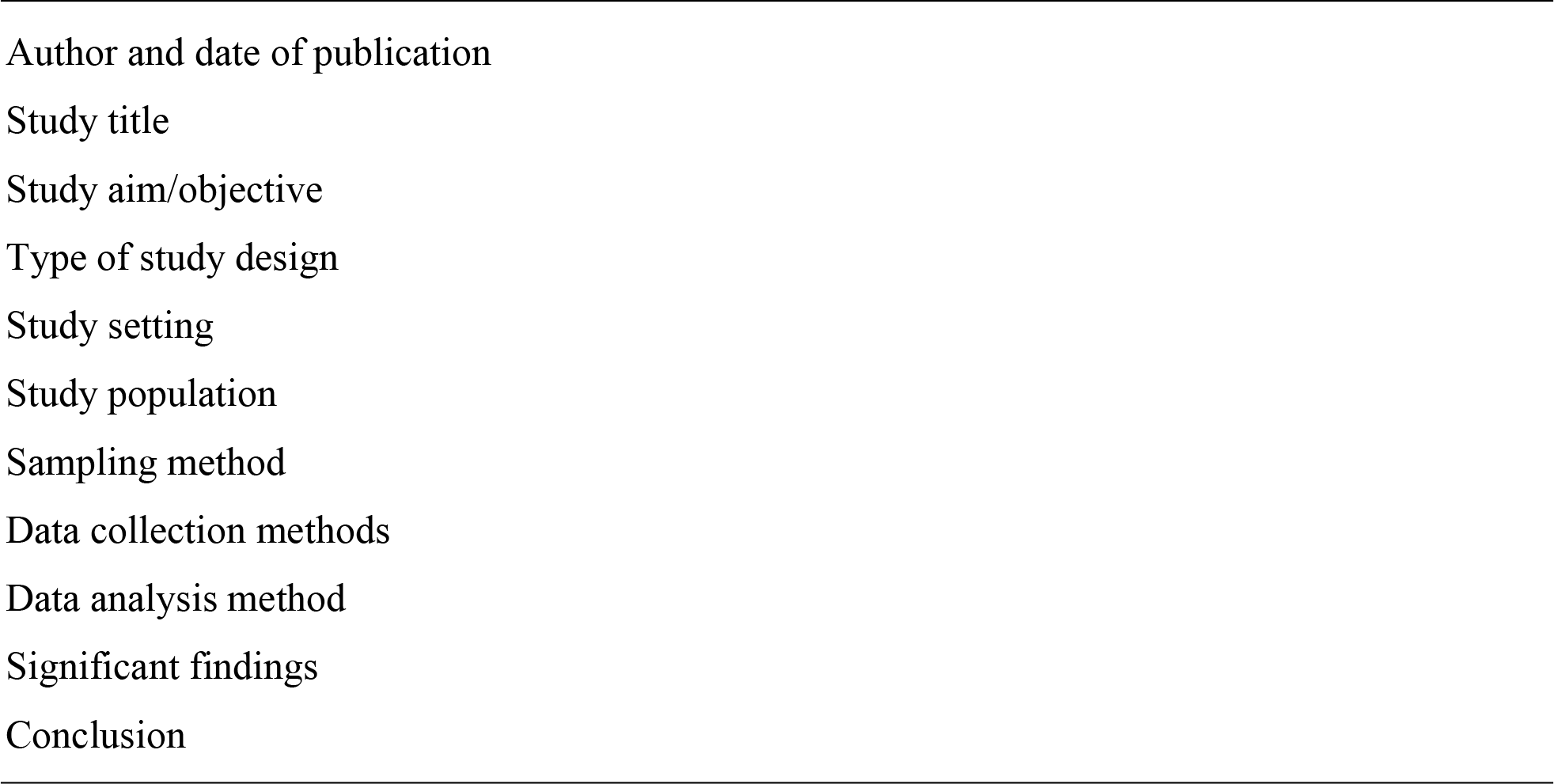
Data extraction form.

## Discussion

The proposed scoping review aims to provide evidence on the existing nurturing care practices for children with NDDs in SSA countries. The review also aims to identify key determinants of care provided to children with NDDs as well as the experiences of the parents/caregivers of children with NDDs. Further, the review will highlight the existing gaps in the provision of nurturing care for children. The review will provide key recommendations on practical best practices and interventions for improving the quality of care and developmental outcomes for children with NDDs. This review also has the potential to contribute to increased awareness of the unmet nurturing care needs of children with NDDs in the region and will provide evidence to assist policymakers and stakeholders in addressing the needs of this vulnerable population.

This review is limited to only SSA countries, and therefore, the scope does not include other LMICs outside SSA. This review may suffer from publication bias, which is why we chose a scoping review approach. The review is also limited to the English language, which may bias the evidence. In addition, the review only considers the main NDDs (ADHD, cerebral palsy, Autism spectrum disorder, learning disorder, communication disorders, and intellectual disorder), excluding all other disabilities, further biasing the evidence.

## Data Availability

Yes

## Author Contributions

**Conceptualization:** Silas Onyango, Patricia Kitsao-Wekulo, **Data curation**: Silas Onyango, Patricia Kitsao-Wekulo, Margaret Nampijja, Linda Oloo, Nelson K. Langat and Kenneth Okelo.

**Formal analysis:** Silas Onyango, Patricia Kitsao-Wekulo, Margaret Nampijja and Kenneth Okelo Paul Otwate, and Nelson K. Langat.

**Methodology:** Silas Onyango, Kenneth Okelo and Margaret Nampijja.

**Project administration:** Silas Onyango, Patricia Kitsao-Wekulo.

**Writing – original draft:** Silas Onyango

**Writing – review & editing:** Silas Onyango, Patricia KitsaoWekulo, Linda Oloo, Kenneth Okelo, Paul Otwate, and Nelson K. Langat.

## References

1. Olusanya BO, Davis AC, Wertlieb D, Boo N-YY, Nair MKCKC, Halpern R, et al. Developmental disabilities among children younger than 5 years in 195 countries and territories, 1990–2016: a systematic analysis for the Global Burden of Disease Study 2016. Lancet Glob Heal. 2018;6(10):1100–21.

2. Olusanya B, Gladstone M, Rukabyarwema JP, Kuper H, Mulaudzi M, Kakooza A, et al. Nurturing care for children with developmental disabilities: a moral imperative for sub-Saharan Africament. 2018;

3. Tamburlini G. Nurturing care for early child development. Vol. 37, Medico e Bambino. 2018. 489 p.

4. WHO. Nurturing Care for Early Childhood Development: A framework for helping children survive and thrive to transform health and human potential. 2018.

5. Black MM, Walker SPS, Fernald LCHL, Andersen CT, DiGirolamo AM, Lu C, et al. Early childhood development coming of age: science through the life course. Lancet [Internet]. 2016;0(0):60–70. Available from: http://linkinghub.elsevier.com/retrieve/pii/S0140673616313897

6. Arksey H, O’Malley L. Scoping studies: Towards a methodological framework. Int J Soc Res Methodol Theory Pract. 2005;8(1):19–32.

7. Levac D, Colquhoun H, O’Brien KK. Scoping studies: Advancing the methodology. Implement Sci. 2010;5(1):1–9.

8. Tricco AC, Lillie E, Zarin W, O’Brien KK, Colquhoun H, Levac D, et al. PRISMA extension for scoping reviews (PRISMA-ScR): Checklist and explanation. Ann Intern Med. 2018;169(7):467–73.

9. Teare G, Taks M. Extending the scoping review framework : a guide for interdisciplinary researchers. Int J Soc Res Methodol. 2019;1–5.

10. Bradbury-Jones C, Aveyard H, Herber OR, Isham L, Taylor J, O’Malley L. Scoping reviews: the PAGER framework for improving the quality of reporting. Int J Soc Res Methodol. 2022;25(4):457–70.

